# Can individualized targets for transcranial magnetic stimulation increase treatment effectiveness in psychiatric disorders? A systematic review and meta-analysis

**DOI:** 10.1101/2021.10.14.21265029

**Authors:** Yanbin Zheng, Zhaojie Zhang, Bo Yang, Weiran Zhou, Xianwei Che, Guang-Heng Dong

**Affiliations:** Centre for Cognition and Brain disorders, School of Clinical Medicine, Hangzhou Normal University, Hangzhou, Zhejiang Province, P.R. China; The Affiliated Hospital of Hangzhou Normal University, Hangzhou, Zhejiang Province, P.R. China; Zhejiang Key Laboratory for Research in Assessment of Cognitive Impairments, Hangzhou, Zhejiang Province, P.R. China

**Keywords:** Noninvasive brain stimulation, Individualized targets, Structural-based methods, Meta-analysis

## Abstract

**Background:** Transcranial magnetic stimulation (TMS) techniques have developed in recent years in research and clinical treatment. The identification of targets for TMS treatment is increasingly individualized based on morphology or function; however, whether individualized TMS targets could increase the treatment effectiveness of psychiatric disorders remains controversial.

**Methods:** A meta-analysis was conducted to explore whether individualized TMS targets are better than standard targets. A total of 3340 studies were identified in a systematic search, and twelve were included in the quantitative review. Among them, eight used a structure-based individualized target selection method, nine were on depression, and four compared unilateral and bilateral stimulant targets.

**Results:** Meta-analyses showed that: (1) individualized TMS targets increased the effectiveness in treating psychiatric disorders; (2) structural-based TMS targets brought additional treatment effectiveness, and PET-based structural selection methods proved to be valid; (3) there was no significant increase in the treatment effects of individualized targets in EEG-based and task-fMRI-based methods; (4) updated stimulant sequences did not increase the individualized target treatment effect; (5) individualized TMS targets showed increased treatment effectiveness in depression but not in schizophrenia; and (6) bilateral stimuli did not show additional effectiveness compared with unilateral stimuli.

**Conclusions:** The current findings revealed that individualized TMS targets show additional treatment effectiveness compared to standard targets in treating psychiatric disorders, and structure-based selection methods are effective in identifying TMS targets. The current conclusions provide directions for future TMS research and provide valuable references for clinicians treating psychiatric disorders.

## Introduction

Noninvasive brain stimulation (NIBS) techniques, such as repetitive transcranial magnetic stimulation (rTMS) or transcranial direct current stimulation (tDCS), have developed in recent years beyond mere clinical application. In rTMS, magnetic fields induce focal electrical currents indirectly and enable focal stimulation of the target area, whereas tDCS involves the alteration of neuronal membrane polarization without triggering action potentials ^1,2^. Although most of these techniques were originally designed for the treatment of neuropsychiatric disorders, they are becoming important approaches in studying the causal relationships between brain functions and behaviors, ^3,4^ and they have also achieved promising results in treating psychiatric disorders ^5,6^, contributing to a deeper understanding of relevant disorders.

Studies have revealed that most psychiatric disorders are accompanied by brain dysfunction and changes in neurotransmitters ^7,8,^ representing an opportunity for devising possible strategies for treating these disorders with NIBS techniques. The effects of rTMS on mood are related to its ability to modulate brain functions, such as cortical excitability ^9^ or blood flow. High-frequency rTMS (>5 Hz) has been shown to increase brain activity both locally and in distant regions, while low-frequency TMS (<1 Hz) can decrease brain activities ^10^. Using various protocols, researchers have achieved valuable progress; for example, Amiaz *et al*. used high-frequency rTMS to target the left dorsolateral prefrontal cortex (DLPFC) to attenuate nicotine cravings through activation effects ^11^. A systematic review suggested that rTMS is a well-tolerated treatment in patients with borderline personality disorder ^12^. Transcranial infrared laser stimulation (TILS) to the DLPFC, which is implicated in emotion regulation, resulted in a context-specific benefit as a monotherapy for reducing fear ^13^. Transcranial alternating current stimulation (tACS) is useful in reducing left frontal alpha oscillations, implying a future treatment strategy for major depressive disorder ^14^.

Choosing stimulation targets is a key step in NIBS studies. Traditionally, researchers targeted one ‘standard’ brain region in all subjects. For example, as studies found that high-frequency rTMS was effective over the left DLPFC ^15^, most studies targeted stimulating the left DLPFC (particularly BA 46 or an area encompassing left BA 9 and BA 46) either using a ‘standard’ procedure that targets an area 5 cm anterior to the hand motor cortical representation ^16,17^ or using neuronavigation devices ^18^. These protocols proved to be more efficacious than sham rTMS in treating psychiatric disorders.

However, the ‘standard’ choice of the target region remains speculative ^19^, and the clinical effects have been only moderate, even when using neuronavigation devices ^18^. First, choosing stimulation targets based on preset cortical brain regions (e.g., the DLPFC) could result in unpredictable and potentially detrimental downstream effects ^20^. Second, the preset target regions have often been challenged by studies; for example, although BA 9 and BA 46 are connected to regions implicated in the regulation of mood, such as the anterior cingulate and the caudate ^21^, it is not clear whether other prefrontal clusters should be targeted. One study suggested that the application of high-frequency rTMS over hypometabolic prefrontal areas would yield better clinical effects ^22^. The standard method to preset a target for all subjects does not consider individual variability.

To overcome these shortcomings, the identification of targets for NIBS treatment is increasingly individualized and based on individual morphology or connectivity. Currently, brain connectivity can be measured at multiple levels, for example, structural connectivity, functional connectivity, and effective connectivity. Structural connectivity refers to the structural integrity of tracts connecting different brain areas; functional connectivity refers to the correlations between region- or source-specific time series; and effective connectivity is the causal influence that neural populations exert over each other. This connectivity could be inferred from functional imaging data; for example, EEG and fMRI are powerful tools to noninvasively probe brain circuits in humans, allowing for the assessment of several cortical properties, such as excitability and connectivity. Individualized NIBS targets have been used in studies, including PET-guided ^23^, EEG-based ^24^ or fMRI-based ^25^ TMS targets.

However, the individualized NIBS targets seem more precise than group-standard targets. Nevertheless, this method has also been challenged by researchers. First, since TMS are not precise and some of the targets are in a rather broad area, a very small target seems unnecessary. Second, we cannot control the subsequent conduct directions after stimulation, and the effect generated in one target could easily be conducted to neighboring brain regions. Third, it has been posited that TMS can impact brain activity in regions that are distant from the stimulation site, and the stimulant, directly or indirectly, propagates to other regions ^26^. Fourth, Ahdab et al. compared the locations of the DLPFC by the 5-cm method of coil positioning and using a neuro-navigation system integrating individual MRI, revealing that the “individualized” procedure failed to accurately locate DLPFC targets, and the navigation methods are costly and time-consuming compared with the estimation method ^27^.

Experimental studies of this issue have also brought controversies. For example, some studies have supported that individualized target are better for treatment effectiveness in psychiatric disorders ^23,28^; in contrast, others have argued that no better effect was observed between individualized targets and standard targets ^29-31^. Thus, a meta-analysis is necessary to elucidate the effects of standard brain regions and individualized brain regions and to establish efficacy and a way to select the best cortical targets for stimulation protocols.

The primary focus of the current study was to conduct a systematic review and meta-analysis to compare the treatment effectiveness of individualized NIBS targets and standard targets in psychiatric disorders. Second, we also wanted to examine the different methods in finding individualized targets to determine which ones are valid and which are not proven, including EEG-based, task-fMRI-based, and structure-based individualized targets. Third, for different psychiatric disorders, individualized NIBS might generate different effects. Fourth, some research has used unilateral individualized NIBS targets, and some has used bilateral targets. We wanted to determine their effectiveness in increasing the treatment effect.

## Methods

### Protocol and registration

This review adhered to the guidelines of the Preferred Reporting Item for Systematic Reviews and Meta-Analysis (PRISMA) ^32,33^. The protocol was registered in the database of the International Prospective Register of Systematic Reviews (PROSPERO, 272000).

### Search strategy

A comprehensive electronic literature search was performed in PubMed, Web of Science, PsycINFO, ProQuest and Embase. The publication date was up to October 2021. The key words used for the search were “depression” OR “schizophrenia” OR “bipolar disorder” OR “substance OR (substance use OR nicotine OR drug OR food OR alcohol)” AND (transcranial OR transcranial magnetic stimulation OR TMS OR TBS) AND (navigated OR navigation OR guided OR individualized OR individual OR personalized).

Two reviewers (YZ and ZZ) independently assessed the titles and abstracts of the initial research results by inclusion criteria and exclusion criteria (see Table 1). Full texts were examined if the abstract information was unclear. Full-texts of potentially eligible studies were then screened. Discrepancies between the two investigators were resolved by consensus. Reference lists of full-text potentially eligible studies and reviews were also checked for missing studies. For missing data, investigators sent emails to authors asking for data.

**Table 1.**
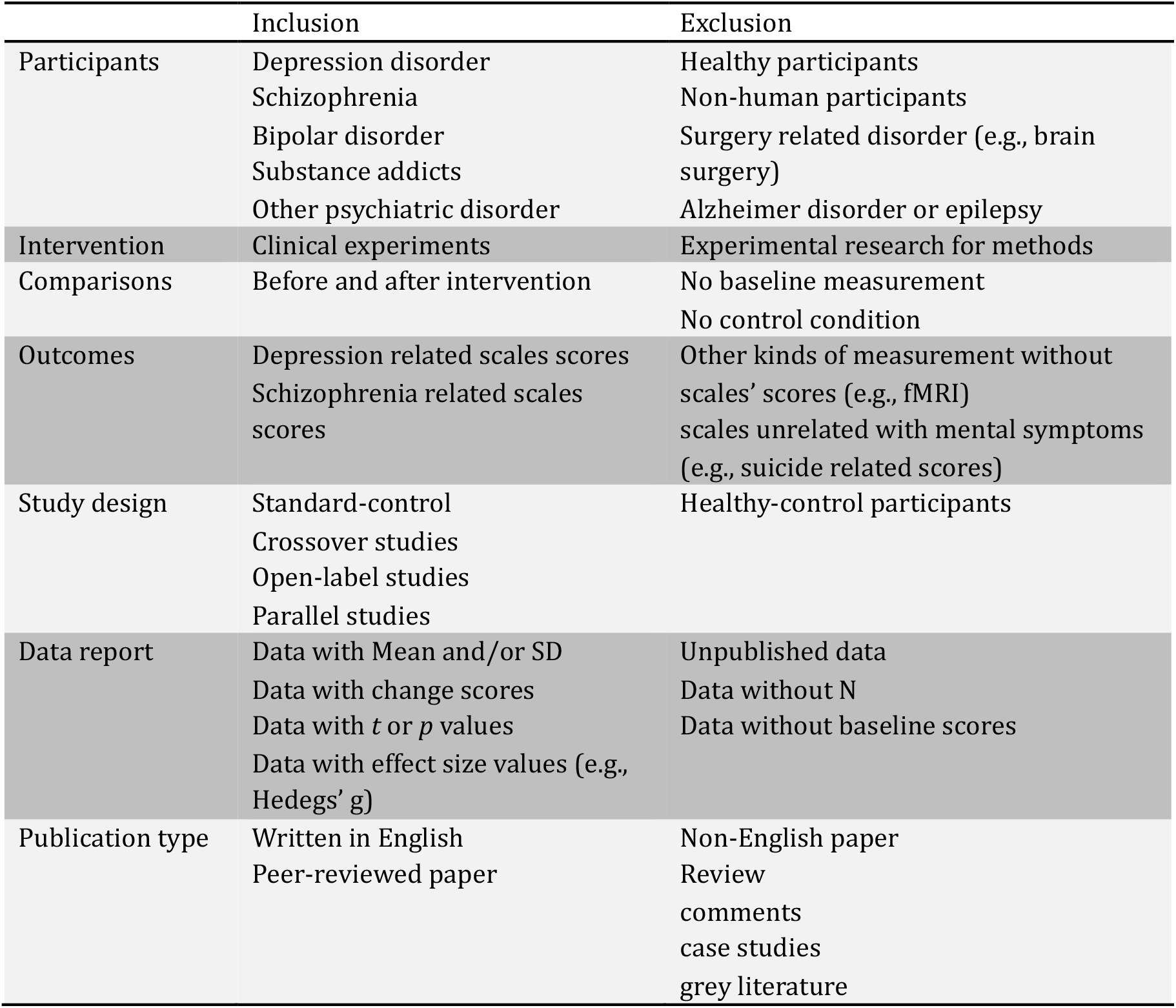
The inclusion and exclusion criteria in current study.

### Selection criteria

The inclusion and exclusion criteria were used to select eligible articles for meta-analysis and systematic reviews, as listed in Table 1. Specifically, the included articles had to satisfy seven criteria: (1) Participants: The participants were psychiatric patients (e.g., depression or schizophrenia); healthy participants or animal experiments were excluded; (2) Intervention: Experiments in searching individualized TMS and clinical research of individualized TMS were included, while methodology of TMS or mechanism studies of TMS were excluded; (3) Comparisons: The scores both before and after measurements were manipulated, but studies were excluded if they lacked baseline measurement and/or control conditions; (4) Outcomes: The scale scores of mental-related scales (e.g., Hamilton depression rating, HAMD) were reported, whereas other types of scores, such as neuroimaging results (e.g., functional magnetic resonance imaging, fMRI) or scores unrelated to mental symptoms (e.g., suicide-related scales), were excluded; (5) Study design: The study design included a standard-control group, crossover group, parallel group and open-label group. The standard control group was defined as the method for locating the target. Concretely, the method of standard grouping for depression refers to use the F5 or F3 as the reference coordinate or a location 5-cm in front of the scalp, for which we called it the standard method as well ^34,35^; (6) Data reporting: The type of data reported was available to compute effect size using Hedge’s g (e.g., mean (M), standard deviation (SD)), but studies were excluded if the data were unavailable or incalculable; (7) Publication type: Peer-reviewed and English-written articles were included.

### Outcome measurement

Most of the participants in our meta-analysis were depression or schizophrenia patients. The majority of depression experiments used depression-related scales to measure the degree of depression. Most experiments used scales including the HAMD ^36,37^, Beck Depression Inventory (BDI) ^38,39^ and Montgomery-Å sberg Depression Scale (MADRS) ^40^. To analyze the results conveniently and uniformly, the result type of the HAMD was chosen as the primary type, and other scales’ scores were translated into the type of HAMD ^41-44^. Thus, we selected HAMD scores as the primary outcome, the BDI scores and MADRS scores were chosen as the secondary outcomes.

Most schizophrenia-related papers used the Auditory Hallucinations Rating Scale (AHRS) or the Positive and Negative Syndrome Scale (PANSS) ^45,46^. Therefore the AHRS scores were selected as the primary scores, and PANSS scores served as the secondary scores when no AHRS scores were assessed. The detailed selecting procedure can be seen in Figure 1.

**Figure 1.**
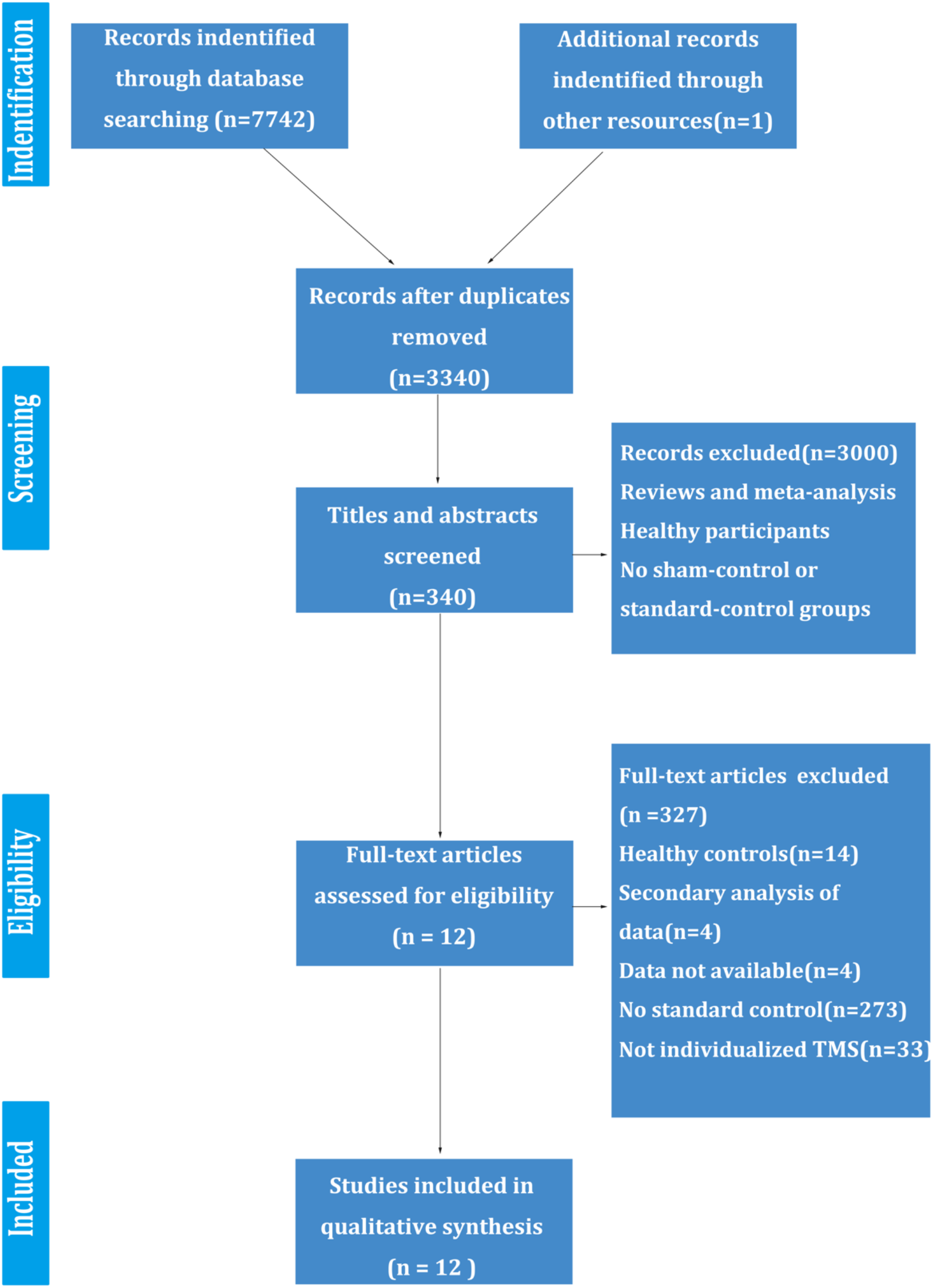
The flow diagram of the meta-analysis.

### Data extraction

Two investigators independently extracted data from the included articles. Sample sizes, change scores, *t* values, *p* values, and M and SD of pre- and postresults were extracted to generate the effect size. Data on the number of participants, mean age of participants, and individualized methods were extracted as well.

### Meta-analysis

All statistical analyses were performed using Comprehensive Meta-analysis Software (version 2).

### Assessment of risk of bias

We assessed the quality of selected studies using the Cochrane Risk of Bias Tool (CRBT) ^47^. In this review, six types of bias were assessed: selection bias, performance bias, detection bias, attrition bias, reporting bias and other bias. Specifically, selection bias included random sequence generation and allocation concealment. Performance bias included the blinding of participants and personnel, and detection bias included the blinding of outcome assessments. Attrition bias and reporting bias were caused by incomplete outcome data and selective reporting, respectively.

### Effect size calculation

The effect size was calculated in three forms. First, M and SD in pre- and postscores and sample sizes of two groups were generated. Second, M and SD of change scores and sample sizes of two groups were manipulated. Finally, the mean of pre- and postscores, *p* values for score changes, and sample sizes were computed. Then, the effect size was calculated with Hedges’s g statistic ^48,^ and significant differences in pooled effect sizes were assessed by 95% confidence intervals (CIs). Since many types of psychiatric disorders were included, Hedges’s g statistic served as a primary indicator to make comparisons.

### Main effect analysis and subgroup analysis

In the main effect analysis, the main effect of individualized TMS and standard TMS was analyzed. In addition, in the subgroup analysis, comparisons between different types of individualized TMS and standard TMS were performed, and different types of diseases were analyzed.

### Heterogeneity test and meta-regression

In systematic reviews and meta-analyses, the conclusions are unclear regarding whether the results varied across studies. Thus, the test of heterogeneity is necessary, which reflects the degree of consistency across studies ^49^. In accordance with other meta-analysis reviews in psychiatric disorders ^44,50^, the Cochrane Q test and I^2^ statistic were used to evaluate significant differences in heterogeneity tests among studies. For the Q test ^51^, *p*<0.05 was considered to indicate significant heterogeneity. The detailed inconsistency levels among studies were assessed by the I^2^ statistic ^52,53^, and an I^2^ index of 0.25, 0.50, and 0.75 indicated small, moderate, and high levels of heterogeneity, respectively. We anticipated the heterogeneity of these studies; therefore, a random-effects model was employed.

Meta regression is another method to search for the original possibilities of heterogeneity ^54^. In this meta-analysis, the number of participants and the mean age of patients were believed to affect the results. Thus, these two variables were entered into the software to calculate the meta regression ^55,56^.

### Publication bias and sensitivity analysis

Publication bias can lead to adverse effects influencing the conclusions of a meta-analysis. Thus, a number of methods have been generated to quantify and analyze publication bias. The funnel plot is an intuitive method to inspect for publication bias ^57^. It provides a graph of effect size against sample size, which is computed as standard error by Hedges’s g statistic ^58^. The distribution position relies on the sample size of the studies. Small-sample size studies will be scattered at the bottom of the funnel plot, while large-sample size studies will be distributed at the top. In addition, a meta-analysis with less publication bias represents a symmetrical scatter in the funnel plot. However, the funnel plot cannot provide specific results, and it is difficult to judge the degree of publication bias. Therefore, Egger’s test and the classic fail-safe N test were used to determine the statistical significance of publication bias. Egger’s test uses a linear regression to examine publication bias ^59^. Then, the classic fail-safe N index further investigates how many missing studies or studies with opposite conclusions are added to the analysis, leading to nonsignificant results in the pooled studies ^60,61^. Moreover, sensitivity analysis is another method to test publication bias, and the one study removal method is commonly used ^62^. The method is removing one study at a time to evaluate whether a single study will influence the results of the meta-analysis.

## Results

Using the selection criteria, all included 12 studies were using TMS. Thus, we narrowed the results from NIBS to TMS.

### Selected studies and characteristics

Online databases identified 17742 records in total (see Figure 1). After duplicated studies were removed, a total of 3340 articles remained. Initial screening of titles and abstracts was performed according to the inclusion and exclusion criteria. In this step, most of the excluded articles were methodology studies. After the screening step was finished, 340 articles remained. In the following manner, reviewers read these articles carefully to further select eligible studies. Thus, a total of 327 articles were excluded for using healthy controls, secondary analysis of data, no standard-control group and unavailable data. Finally, only 12 articles were included in the meta-analysis and systematic review. Demographic information and other characteristics of the articles are shown in Table 2.

**Table 2.**
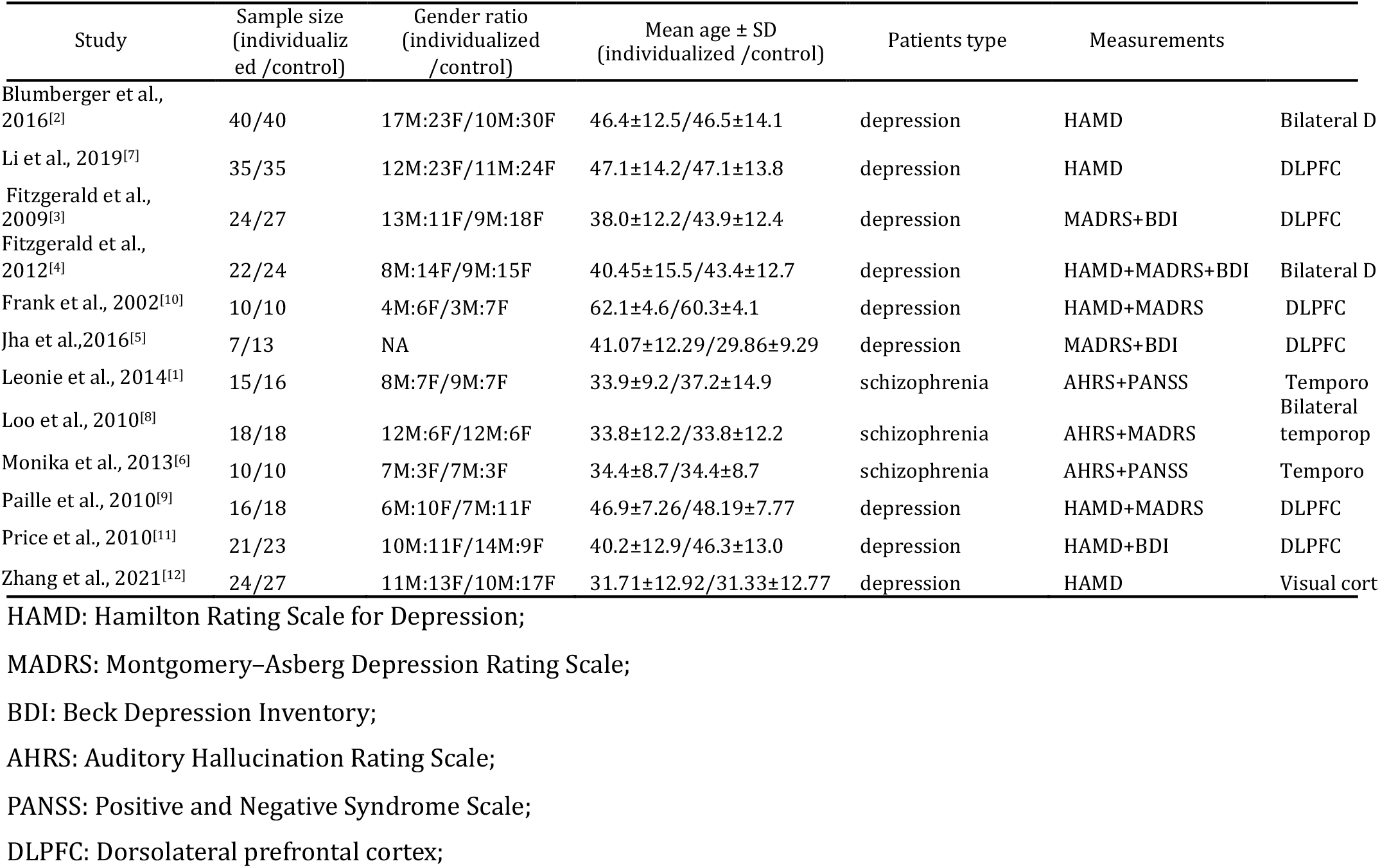
The characteristics of included studies.

A range of individualized TMS method were investigated combined with other equipment, like EEG (n=1), PET (n=3) or MRI (n=5). The types of psychiatric disorders were basically depression (n=9) and schizophrenia (n=3). The TMS treatment target of DLPFC was basically selected in depression while the temporal region was elected as the TMS treatment target of schizophrenia. The total pulses of TMS almost no less than 16000 pluses. The HAMD scales and AHRS scales were the primary outcome measures to evaluate the symptoms of depression and schizophrenia, which in accordance with previous studies^63,64^.

Besides, the quality of these articles was assessed by CRBT (See Figure 2). In specific, low risks of selection bias indicated that randomized allocation in different group while high risks of selection bias included pseudorandom allocation or participants knew that what kinds of treatment they received. The low risks of performance bias were usually double-blind design or experimenters would not affect the results. During low risks of detection bias, it suggested that researchers used proper methods or scales to measure the symptoms of disorders without controversy. As for attrition bias and reporting bias, low risks of the two items indicated that researchers did not report the results by selecting elaborately leading to the results perfect. Most of the selected studies had low or median risk, indicating that the selected articles were basically low risk.

**Figure 2.**
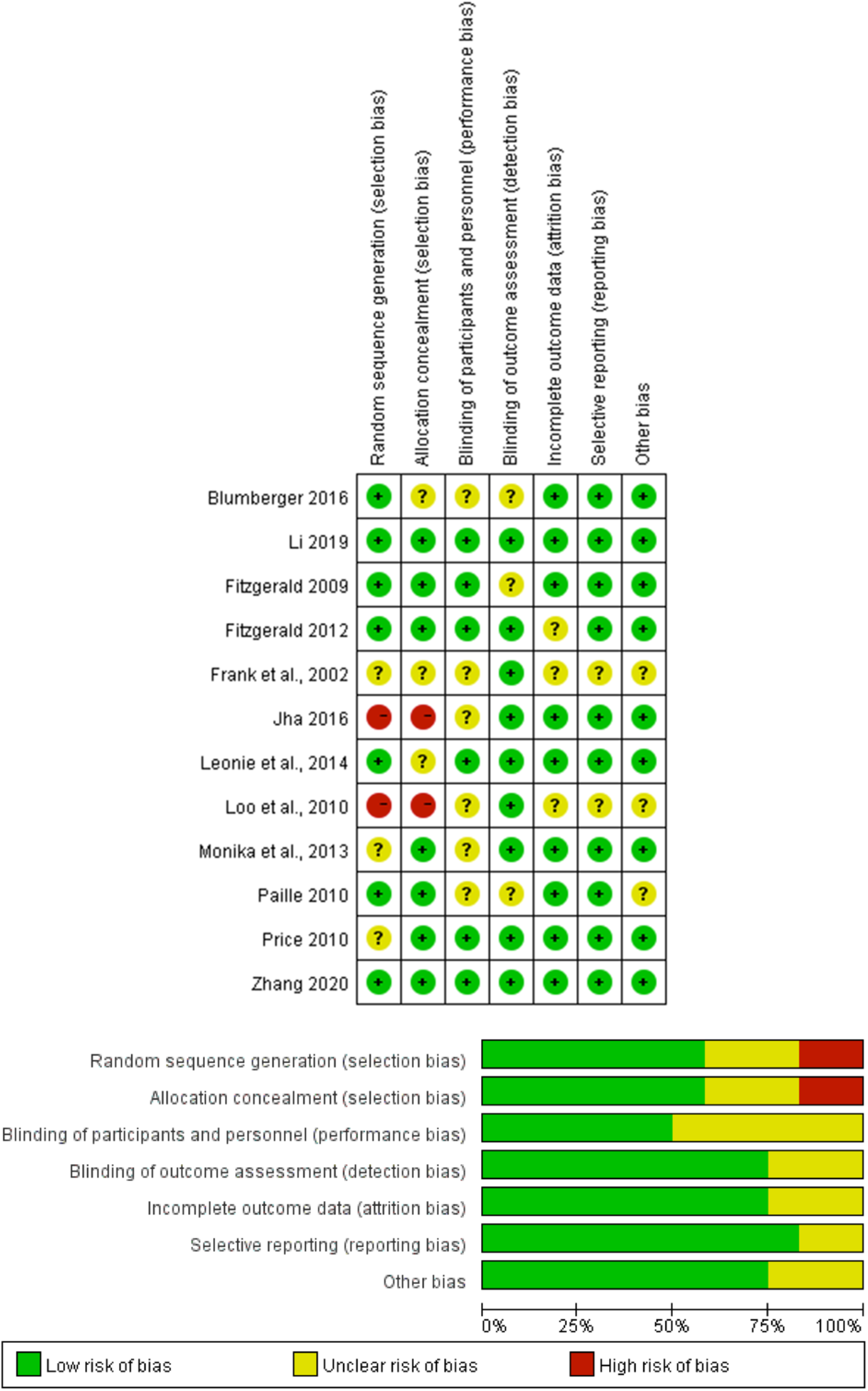
Risk of bias evaluation of selected studies. The results assessed by Cochrane Risk of Bias Tool. It concluded six parts, which are selection bias, performance bias, detection bias, attrition bias, reporting bias and other bias. The results suggested that almost more than fifty percentage in each kind of bias was low.

### Results of publication bias and sensitivity analysis

Funnel plot inspection showed a basically symmetric arrangement mainly located at the middle or top, suggesting that no significant publication bias was found. Egger’s test (*t*_11_=1.609, *p*=0.13, two-tailed) confirmed the results. In addition, the classic fail-safe N index indicated 29 missing studies that would bring *p*>0.05 for the meta-analysis (Figure 2).

Subsequently, the results of removing one study suggested that whichever study was removed, the results remained significant. In particular, no study significantly affected the meta-analysis results between individualized TMS and standard TMS.

### Meta-analysis of individualized TMS stimulant targets and standard targets

A random-effect meta-analysis of the extracted 12 articles encompassed 489 individuals with psychiatric disorders, with 240 patients with psychiatric disorders receiving individualized TMS treatments and another 249 patients with psychiatric disorders cured with standard TMS treatments. The results showed that the individualized TMS treatments were notably better than the standard treatments (Hedges’s g=0.300; 95% CI= [0.128, 0.473]; *p*=0.001) (see Figure 3).

**Figure 3.**
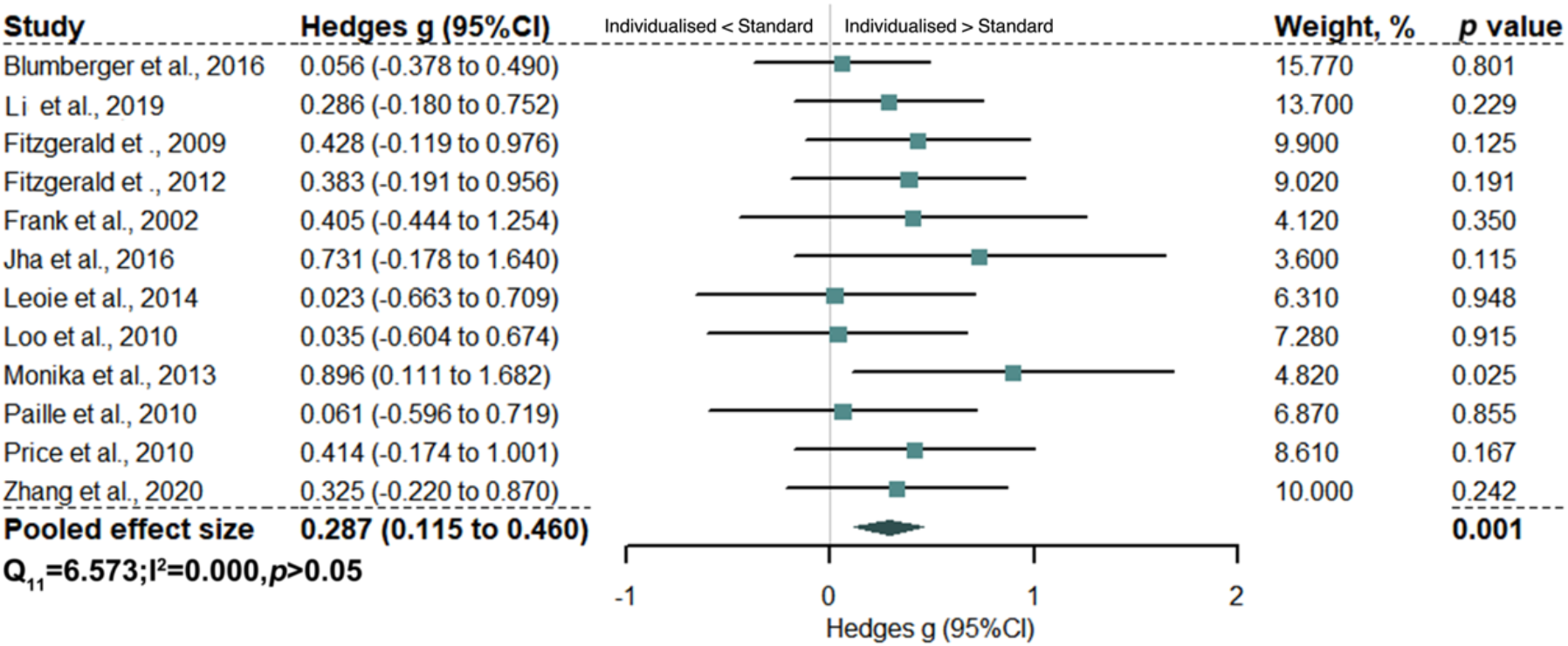
Main effect results between individualized targets and standard ones. The size of squares was proportional to study weights. The rhombus marker indicated that pooled effect size.

### Subgroup analysis of the effectiveness using structural-based methods in selecting individualized TMS targets

The random-effect meta-analysis method was employed in the subgroup analysis as well. Our results found that the types of individualized TMS were divided into six forms: electroencephalography (EEG)-guided, positron emission tomography (PET)-guided, structure magnetic resonance imaging (sMRI)-guided, task fMRI-guided, bilateral or contralateral guidance and sequence individualized. Specifically, the method of EEG-guided stimulation uses EEG for precisely location ^65^. PET-guided and sMRI-guided methods are also called structure-guided methods using neuroimaging by PET or MRI ^66,67^. Task-fMRI guidance asked participants to finish some tasks during fMRI so that the target could be precisely located according to their brain responses while finishing tasks ^68^. Since standard TMS in major depression disorder usually chooses the DLPFC as the stimulation target to treat depression, the left temporal regions of schizophrenia were basically selected as standard targets. The bilateral- or contralateral-guided method used two TMS machines to stimulate the participants or one machine to stimulate the contralateral regions of participants. ^35,46^. Sequence-individualized methods modify the sequences of TMS to treat psychiatric disorders ^34,69^.

As shown in Figure 4A, the results of structure-guided individualized TMS targets yielded an additional treatment effect compared to standard targets (Hedges’s g=0.344; 95% CI= [0.100, 0.588]; *p*=0.006). Further analyses showed that the PET-guided TMS targets significantly increased the efficacy (Hedges’s g=0.482; 95% CI= [0.041, 0.923]; *p*=0.032) (Figure 4B), and structural MRI-guided individualized targets were marginally significant compared with standard targets (Hedges’s g=0.283; 95% CI= [-0.010, 0.577]; *p*=0.058) (Figure 4C).

**Figure 4.**
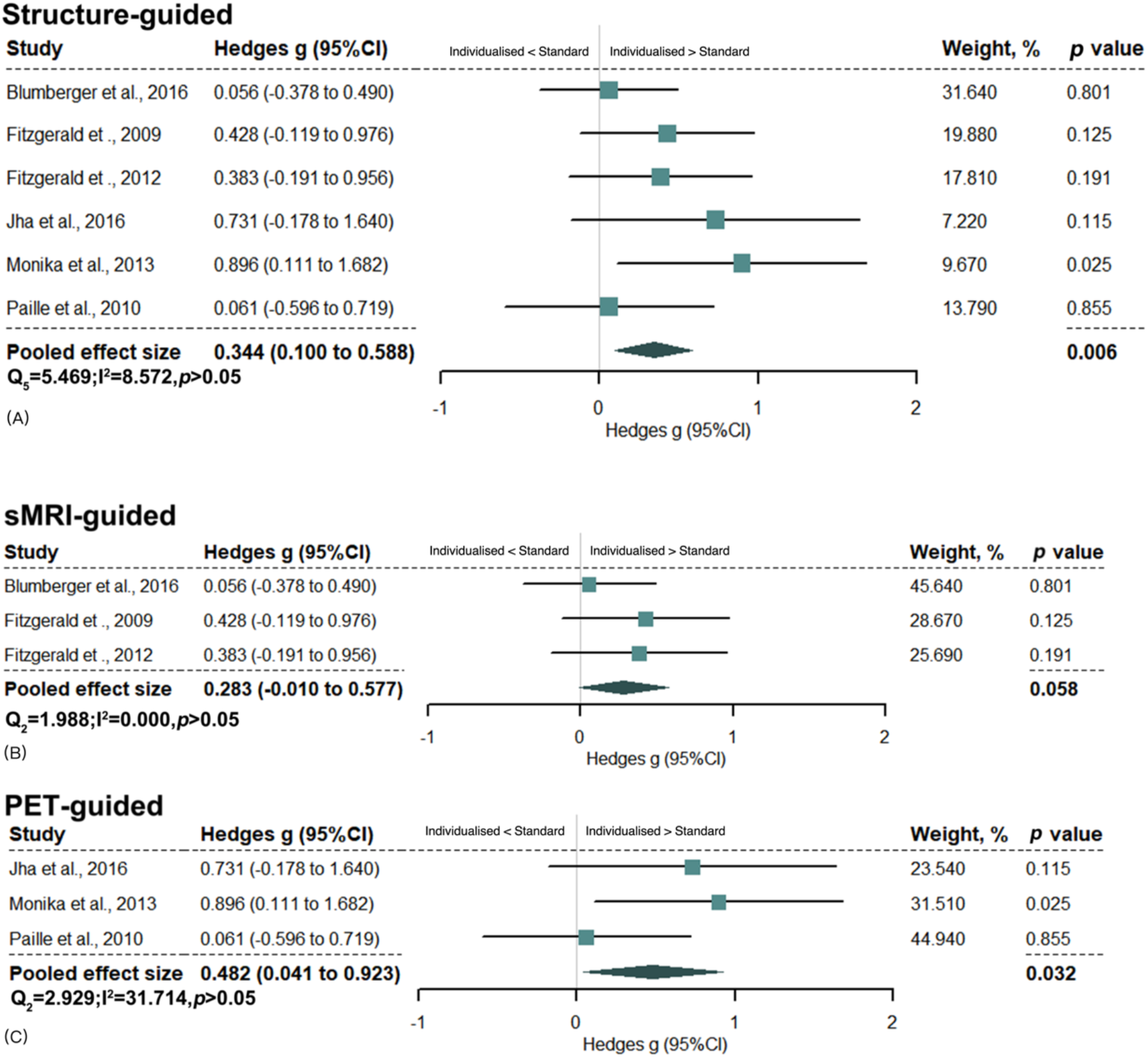
Effect results of the structure-guided subgroup. The subgroup of the structure-guided individualized method (A). Structural MRI (sMRI)-guided individualized method (B), and PET guided individualized method (C). The size of squares was proportional to study weights. The rhombus marker indicated that pooled effect size.

### Subgroup analysis of the effectiveness using unilateral and bilateral individualized TMS targets

Regarding the lateral issue, the results of the bilateral-or-contralateral-stimulant method did not show additional effectiveness compared with standard target therapy (Hedges’s g=0.155; 95% CI= [-0.123, 0.434]; *p*=0.274) (Figure 5A).

**Figure 5.**
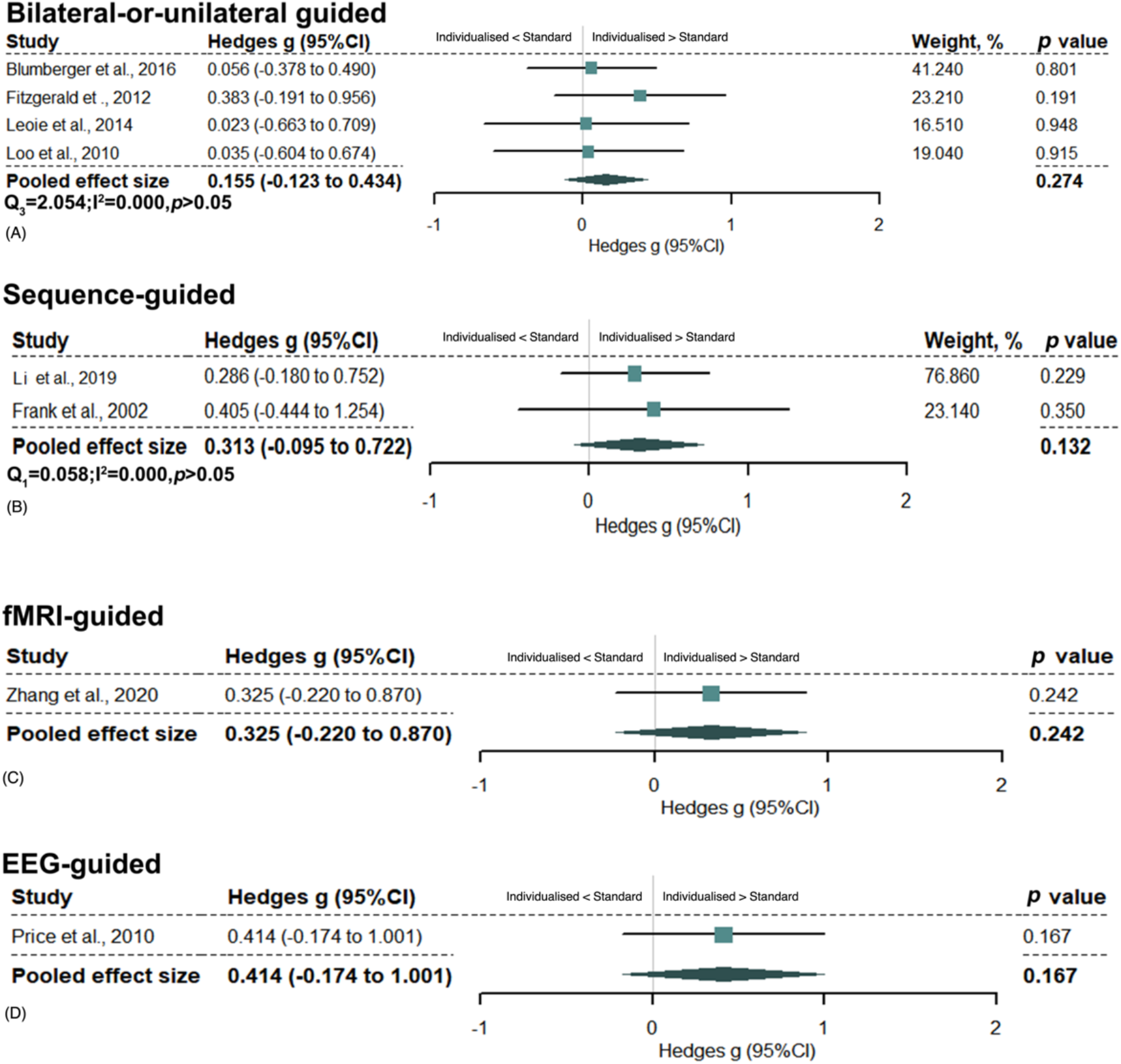
Subgroup analysis of individualized methods. Subgroup results of individualized TMS by its types. Bilateral-or-unilateral guided individualized methods (A); The sequence-guided individualized methods (B); The task fMRI-guided individualized method (C); The EEG-guided individualized method (D). The size of squares was proportional to study weights. The rhombus marker indicated that pooled effect size.

### Subgroup analysis of the effectiveness of stimulant sequences using individualized TMS targets

The upgraded stimulant sequence for individualized TMS targets did not generate additional treatment efficacy compared with standard ones (Hedges’s g=0.313; 95% CI= [-0.095, 0.722]; *p*=0.132) (Figure 5B)

### Subgroup analysis of the effectiveness using task-fMRI and EEG in selecting individualized TMS targets

The the task-fMRI-based target (Hedges’s g=0.325; 95%CI= [-0.220, 0.870]; *p*=0.242) (Figure 5C) and EEG-guided TMS target (Hedges’s g=0.414; 95%CI= [-0.174, 1.001]; *p*=0.167) (Figure 5D) did not generate additional treatment efficacy in treating psychiatric disorders compared to standard targets.

### Subgroup analysis of the effectiveness in different psychiatric disorders using individualized TMS targets

We also performed a subgroup analysis of different psychiatric disorders. The results showed that the individualized TMS methods increased the treatment effectiveness in depression patients (Hedges’s g=0.310; 95% CI= [0.119, 0.501]; *p*=0.001) (Figure 6A) but not in schizophrenia patients (Hedges’s g=0.256; 95% CI= [-0.146, 0.658]; *p*=0.211) (Figure 6B).

**Figure 6.**
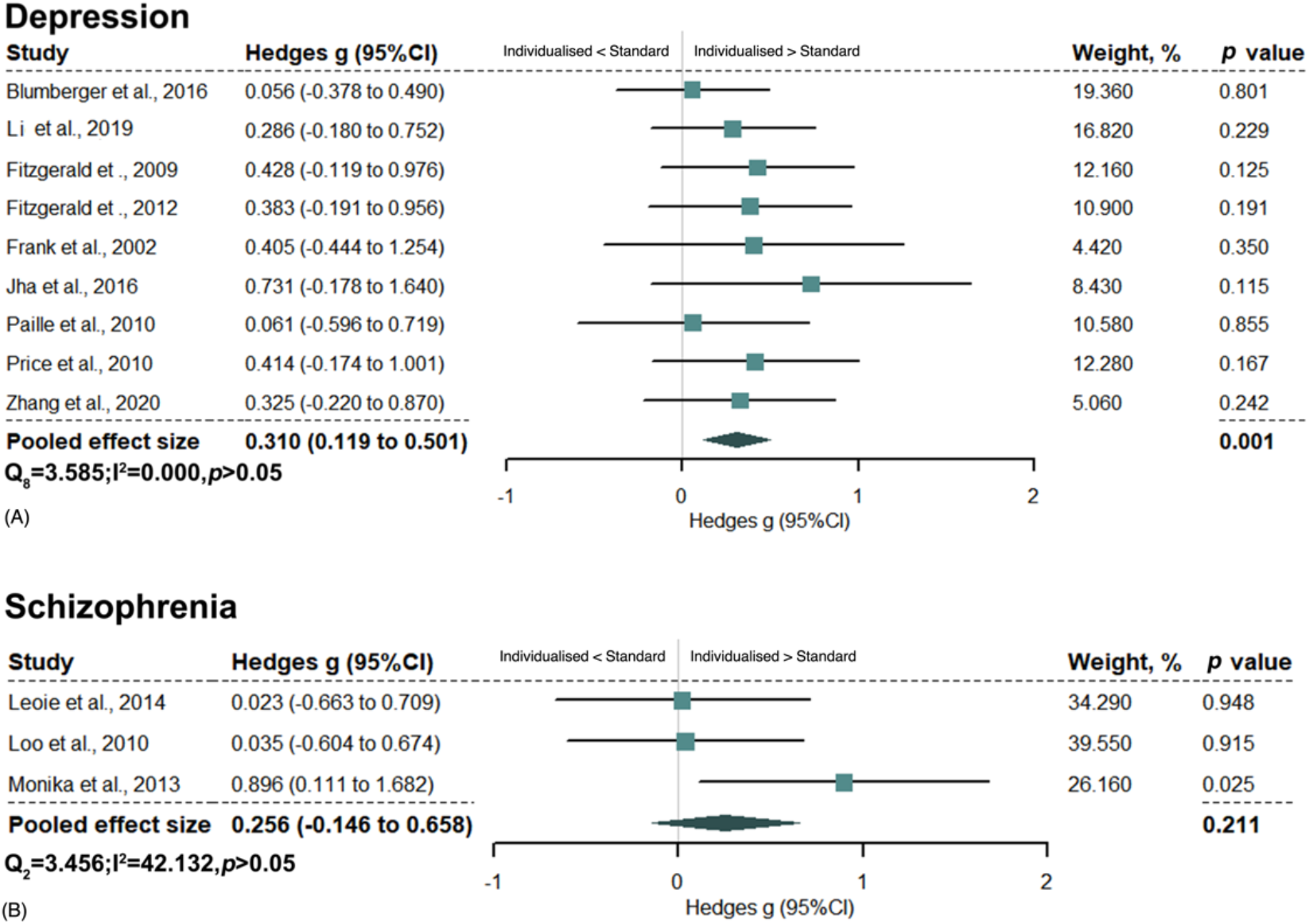
Subgroup analysis of patient types in individualized methods. Subgroup results of individualized methods through patient types. It included depression (A) and schizophrenia (B) patients. The size of squares was proportional to study weights. The rhombus marker indicated that pooled effect size.

### Results of heterogeneity test and meta-regression

During the heterogeneity testing of all articles, nonsignificant heterogeneity among studies was illuminated in the meta-analysis (Q_11_=7.098, I^2^=0.000, *p*=0.791). Subsequently, the mean age of participants (Figure 7A), the total pulses (Figure 7B), and the number of participants (Figure 7C) and were used as the variables in the meta-regression test. The sample sizes of the studies were significantly negatively correlated with effect sizes (intercept=0.546, *p*=0.029), but the mean age of the participants showed no significance (intercept=0.566, *p*=0.286).

**Figure 7.**
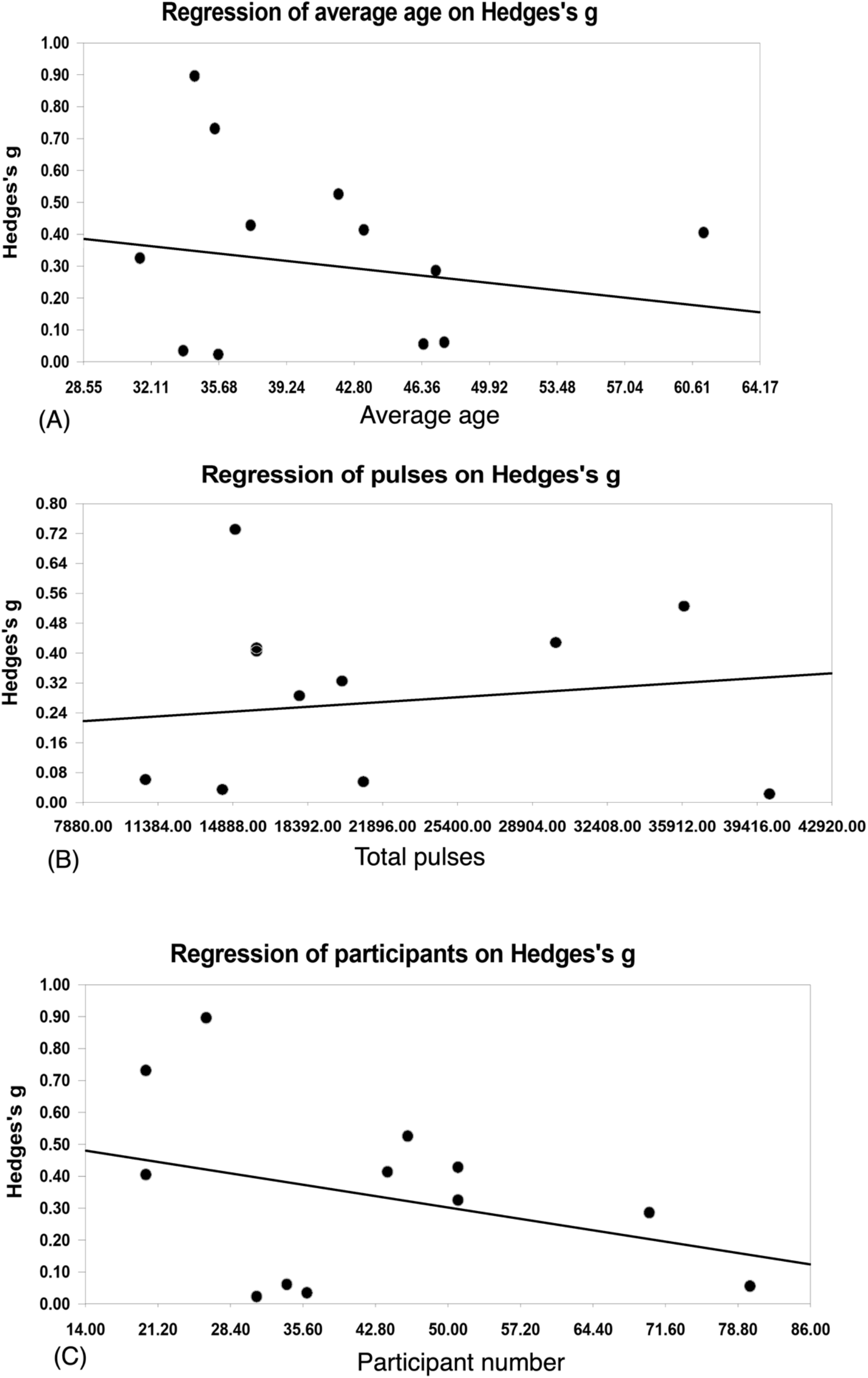
Results of meta regression among three variables. The figure (A) is the results of meta regression of average age of overall participants. The linearity in the figure indicated that negative correlation showed between average age and meta results. The circles indicated average age in each included article. The figure (B) suggested that the results of total TMS pulses. The linearity in the figure indicated that positive correlation showed between total TMS pulses and meta results. The circles indicated total TMS pulses in each included article. The figure (C) is the results of number of participants. The linearity in the figure indicated that negative correlation showed between number of participants and meta results. The circles indicated number of participants in each included article.

## Discussion

The present study explored the effectiveness of individualized TMS stimulant targets compared to ‘standard’ targets. The results showed that the individualized targets increased the treatment effectiveness compared with standard targets in treating psychiatric disorders. Further analyses of subgroups were also performed.

### Individualized TMS targets increased stimulant effectiveness compared with standard targets

The meta-analyses found that individualized TMS targets were more effective than standard targets in treating psychiatric disorders. Studies have posited that TMS can impact brain activity in regions that are distant from the stimulation site and modulate functional connectivity between pairs of brain regions that are not directly stimulated ^26^. The focuses in the psychiatric groups are usually deep in the brain but not in the cerebral cortex; thus, stimulation using traditional ‘standard’ TMS targets might be less effective in reaching the focus of the disordered deep brain, hindering the treatment effectiveness of TMS. In contrast, connectivity-based stimulant targets provide the possibility that transfer the cortex-based stimulation to the deep brain targets ^70^, generating a better treatment effect than standard targets. However, all of these explanations are speculative, and more studies are needed to clarify the potential mechanism underlying this process.

Not all studies agreed with the meta-analysis results that individualized TMS targets are better than standard targets. They argued that no difference was observed between the 5-cm method of coil positioning and individualized target by MRI-based targeting to find the DLPFC ^27^. Potential reasons might include the target brain regions, such as the DLPFC, being affected in psychiatric disorders or covering a relatively large area and there being no certainty regarding which part of it is likely to be critically involved in this disorder. In addition, there are many ways to select individualized TMS targets, such as EEG-based and task-based targets, which might be reasons for the negative results of these studies.

### Structural-based individualized TMS targets increased treatment effectiveness

We further divided the data into groups according to the methods for selecting targets that they used. First, the structure-based individualized TMS target showed significant efficacy in increasing the effectiveness in treating psychiatric disorders compared with standard targets. This outcome occurred perhaps because the structural-based selection methods have the advantage that they could provide a precise target compared to standard 5-cm methods. This thinking is in line with the explanations that the targeted brain regions are relatively large, and a simple 5-cm method only provided estimated targets of the brain region; however, since the brain monography features vary across subjects, the 5-cm methods can only find the approximate location but not a precise location. Structure-based targets could overcome these shortcomings and provide a rather precise location for stimuli, which might be why the individualized TMS target increased the effectiveness of treatment in psychiatric disorders.

There are two types of structural-based targets: PET-based and MRI-based. We performed further analyses to find their effectiveness separately. The results showed that PET-based selection methods showed significant additional effects and that MRI-based methods were marginally significant. PET offers an option to identify the cerebral cortical area of metabolic derangement for stimulation and is a scientific way to administer TMS at the site. It is a potential tool for investigating regional cerebral blood flow in a range of psychiatric disorders ^71^ and hence can be used to localize the site of stimulation for TMS. Conversely, the MRI-based target selection method could only provide a precise target on monography, which might be why it is not accurate as PET-based results.

### Individualized TMS targets increased the treatment effectiveness in depression but not in schizophrenia

Of the twelve publications, nine of them were on depression, and three were on schizophrenia. Thus, we divided the studies into subgroups according to the types of psychiatric disorders. The results showed that the individualized TMS targets increased treatment effectiveness in depression patients but not in schizophrenia patients. Patients with baseline hypometabolism in the prefrontal cortex have been reported to show a better response to high-frequency rTMS stimulation ^72^. High-frequency rTMS could play a role in increasing regional cerebral blood flow, which is associated with improvement in depressive symptoms. The individualized targets provided a precise target for high-frequency rTMS to increase the regional cerebral blood flow and thus improved the treatment effectiveness.

For schizophrenia patients, all of the studies used individualized TMS targets to treat verbal or auditory hallucinations; however, no additional effectiveness was observed in these studies. One reason for the negative finding is that hallucinations, unlike depression, do not have a direct connection with regional cerebral blood flow, and the precise location might not generate an additional treatment effect. Another reason might be that the scale used to measure hallucinations was not sufficiently sensitive to capture the individual nature of the changes experienced by each subject ^73^.

### Bilateral TMS target stimuli did not show additional effectiveness compared with unilateral TMS target stimuli

Among these studies, four studies compared bilateral stimulant sites and unilateral stimuli in treating psychiatric disorders. The meta-analysis of these studies showed that there is a consistent pattern of improved response in unilateral, compared to bilateral, treatment (both superior to sham); however, there is no difference between these two methods. Although studies have shown a robust, superior response in bilateral treatment compared to sham treatment ^74,75^, in direct comparisons, the meta-results showed that bilateral stimuli present a similar response (even inferior) compared to unilateral treatment. This outcome suggests that bilateral brain stimulation is not a factor influencing the TMS outcome, and finding a scientific way to localize a precise target is more important than trying bilateral targets in TMS treatment.

### No additional effect was observed in EEG-based, task-fMRI-based TMS individualized target methods or in updated stimulant sequences

In addition to the structure-based targets, no additional effectiveness in treatment was observed in EEG-guided targets and task-fMRI-based targets. The same feature of these studies is that they were all functionally based selection methods. Studies have shown that functional-based brain locations vary across different studies and different measures in one subject ^76,77^. That is, functionally based individualizing methods (functional connectivity, effective connectivity; brain regions responsible for specific tasks) involved in the first measure might not be observed in another measure, making it difficult to find a solid target to stimulate. Future studies should find ways to render the methods replicable in different measures, which could explain the nonsignificant effect of an increase in the treatment of psychiatric disorders. Another reason for the insignificant effect of the individualized targets is the limited number of studies using functional methods, and more studies are needed to draw scientific conclusions.

In addition to these endeavors to find precise stimulant targets to improve the effectiveness of treatment, one study attempted to use prolonged intermittent theta burst stimulation with a triple dose of the standard protocol to improve the effectiveness of treatment ^29^. That study found that left prefrontal prolonged intermittent theta burst stimulation monotherapy is effective for the treatment of recurrent depression; however, the MRI-guided method of coil targeting is not better than the standard method. The results suggest that the anti-psychiatric disorder mechanisms are still not exhaustively understood, and they might involve mechanisms not associated with the locus of stimulation.

### Risk of bias and publication bias

Risk of bias can affect the results of a meta-analysis. Our data indicated that these included studies tended to be adequate in the generation of random sequences, double blinded studies and reports of outcome data. Studies have found that age could predict low-frequency transcranial magnetic stimulation efficacy ^78,79^. In the current study, we performed regression analyses and found that age was not a factor that affected the final conclusion. Our study proved that the sample size could affect the final results, suggesting that future studies should attempt to include larger sample sizes. In summary, there was no clear evidence of publication bias in the current dataset. The current conclusions are solid and have a good representation for current studies.

### Limitations and future directions

Several limitations should be mentioned. First, we were intended to explore all kinds of individualized NIBS target in treating psychiatric disorders, however, after selecting procedure, only TMS studies were finally included in current study (Few studies trying individualized stimulant targets using other stimulate techniques). Thus, we narrowed the conclusions to TMS; Second, functional-based targets should be promising locations in TMS treatment. However, there have been only a limited number of publications on EEG-based, task-fMRI-based TMS targets. Third, the development of new TMS interventions should consider the propagation of stimulation effects beyond the immediate stimulation vicinity. The idea of the ability of pulse propagation being limited to “one synapsis away” has been progressively dismissed in favor of more extended network propagation. Fourth, all of the studies included only short-term effects, and future studies should test the effects of individualized targets with larger sample sizes and longer-term follow-ups.

## Conclusions

Based on these analyses, we can draw the following conclusions. First, the current study found that individualized TMS targets increased effectiveness in treating psychiatric disorders, suggesting an important method for clinical treatment. Second, structural-based TMS target selections yielded additional treatment effectiveness, and PET-based structural selection showed significant results. Third, there was no significant increase in the treatment efficacy of individualized targets in EEG-based and task-fMRI-based methods or updated stimulant sequences. Fourth, individualized TMS targets showed additional effectiveness in depression but not in schizophrenia. Fifth, bilateral stimulant targets did not generate additional treatment effectiveness compared with unilateral targets.

## Data Availability

All data produced in the present study are available upon reasonable request to the authors
http://QuickConnect.cn/others
ID: guests
PIN

## Acknowledgements

This research was supported by The Cultivation Project of Province-levelled Preponderant Characteristic Discipline of Hangzhou Normal University (20JYXK008), and Zhejiang Provincial Natural Science Foundation (LY20C090005). The funding agencies did not contribute to the experimental design or conclusions, and the views presented in the manuscript are those of the authors and may not reflect those of the funding agencies. This article has been posted on the preprint server BioRxiv.

## Author contribution

Yanbin Zheng initialized this research and screened the publications, analyzed the data, prepared the figures, tables and wrote the methods and results sections. Zhaojie Zhang, Bo Yang, and Weiran Zhou screened the publications; Xianwei Che helped in meta-analyses method. Guang-Heng Dong designed this research and edited the manuscript. All authors contributed to and approved the final manuscript.

## Data Availability

The data stored at our lab-based network attachment system: http://QuickConnect.cn/others. ID:guests; PIN

## References

1. Wing VC, Barr MS, Wass CE, et al. Brain stimulation methods to treat tobacco addiction. Brain Stimul. 2013;6(3):221–230.

2. Tseng PT, Jeng JS, Zeng BS, et al. Efficacy of non-invasive brain stimulation interventions in reducing smoking frequency in patients with nicotine dependence: a systematic review and network meta-analysis of randomized controlled trials. Addiction. 2021.

3. Kozyrev V, Staadt R, Eysel UT, Jancke D. TMS-induced neuronal plasticity enables targeted remodeling of visual cortical maps. Proc Natl Acad Sci U S A. 2018;115(25):6476–6481.

4. Thut G, Veniero D, Romei V, Miniussi C, Schyns P, Gross J. Rhythmic TMS causes local entrainment of natural oscillatory signatures. Curr Biol. 2011;21(14):1176–1185.

5. Smith RC, Boules S, Mattiuz S, et al. Effects of transcranial direct current stimulation (tDCS) on cognition, symptoms, and smoking in schizophrenia: A randomized controlled study. Schizophr Res. 2015;168(1-2):260–266.

6. Trojak B, Meille V, Achab S, et al. Transcranial Magnetic Stimulation Combined With Nicotine Replacement Therapy for Smoking Cessation: A Randomized Controlled Trial. Brain Stimulation. 2015;8(6):1168–1174.

7. Perera T, George MS, Grammer G, Janicak PG, Pascual-Leone A, Wirecki TS. The Clinical TMS Society Consensus Review and Treatment Recommendations for TMS Therapy for Major Depressive Disorder. Brain Stimul. 2016;9(3):336–346.

8. Philip NS, Barredo J, Aiken E, et al. Theta-Burst Transcranial Magnetic Stimulation for Posttraumatic Stress Disorder. Am J Psychiatry. 2019;176(11):939–948.

9. Pascual-Leone A, Valls-Sole J, Brasil-Neto JP, Cohen LG, Hallett M. Akinesia in Parkinson’s disease. I. Shortening of simple reaction time with focal, single-pulse transcranial magnetic stimulation. Neurology. 1994;44(5):884–891.

10. Speer AM, Kimbrell TA, Wassermann EM, et al. Opposite effects of high and low frequency rTMS on regional brain activity in depressed patients. Biol Psychiatry. 2000;48(12):1133–1141.

11. Amiaz R, Levy D, Vainiger D, Grunhaus L, Zangen A. Repeated high-frequency transcranial magnetic stimulation over the dorsolateral prefrontal cortex reduces cigarette craving and consumption. Addiction. 2009;104(4):653–660.

12. Konstantinou GN, Trevizol AP, Downar J, et al. Repetitive transcranial magnetic stimulation in patients with borderline personality disorder: A systematic review. Psychiatry Res. 2021;304:114145.

13. Zaizar ED, Papini S, Gonzalez-Lima F, Telch MJ. Singular and combined effects of transcranial infrared laser stimulation and exposure therapy on pathological fear: a randomized clinical trial. Psychol Med. 2021:1–10.

14. Riddle J, Alexander ML, Schiller CE, Rubinow DR, Frohlich F. Reduction in left frontal alpha oscillations by transcranial alternating current stimulation in major depressive disorder is context-dependent in a randomized clinical trial. Biol Psychiatry Cogn Neurosci Neuroimaging. 2021.

15. Pascual-Leone A, Rubio B, Pallardo F, Catala MD. Rapid-rate transcranial magnetic stimulation of left dorsolateral prefrontal cortex in drug-resistant depression. Lancet. 1996;348(9022):233–237.

16. George MS, Lisanby SH, Avery D, et al. Daily left prefrontal transcranial magnetic stimulation therapy for major depressive disorder: a sham-controlled randomized trial. Arch Gen Psychiatry. 2010;67(5):507–516.

17. Paus T, Barrett J. Transcranial magnetic stimulation (TMS) of the human frontal cortex: implications for repetitive TMS treatment of depression. J Psychiatry Neurosci. 2004;29(4):268–279.

18. Herwig U, Lampe Y, Juengling FD, et al. Add-on rTMS for treatment of depression: a pilot study using stereotaxic coil-navigation according to PET data. Journal of Psychiatric Research. 2003;37(4):267–275.

19. Fitzgerald PB, Huntsman S, Gunewardene R, Kulkarni J, Daskalakis ZJ. A randomized trial of low-frequency right-prefrontal-cortex transcranial magnetic stimulation as augmentation in treatment-resistant major depression. Int J Neuropsychopharmacol. 2006;9(6):655–666.

20. Cocchi L, Zalesky A, Nott Z, Whybird G, Fitzgerald PB, Breakspear M. Transcranial magnetic stimulation in obsessive-compulsive disorder: A focus on network mechanisms and state dependence. Neuroimage Clin. 2018;19:661–674.

21. Barrett J, Della-Maggiore V, Chouinard PA, Paus T. Mechanisms of action underlying the effect of repetitive transcranial magnetic stimulation on mood: behavioral and brain imaging studies. Neuropsychopharmacology. 2004;29(6):1172–1189.

22. Kimbrell TA, Little JT, Dunn RT, et al. Frequency dependence of antidepressant response to left prefrontal repetitive transcranial magnetic stimulation (rTMS) as a function of baseline cerebral glucose metabolism. Biological Psychiatry. 1999;46(12):1603–1613.

23. Jha S, Chadda RK, Kumar N, Bal CS. Brain SPECT guided repetitive transcranial magnetic stimulation (rTMS) in treatment resistant major depressive disorder. Asian J Psychiatr. 2016;21:1–6.

24. Price GW, Lee JW, Garvey CA, Gibson N. The use of background EEG activity to determine stimulus timing as a means of improving rTMS efficacy in the treatment of depression: a controlled comparison with standard techniques. Brain Stimul. 2010;3(3):140–152.

25. Zhang Z, Zhang H, Xie CM, et al. Task-related functional magnetic resonance imaging-based neuronavigation for the treatment of depression by individualized repetitive transcranial magnetic stimulation of the visual cortex. Sci China Life Sci. 2021;64(1):96–106.

26. Vink JJT, Mandija S, Petrov PI, van den Berg CAT, Sommer IEC, Neggers SFW. A novel concurrent TMS-fMRI method to reveal propagation patterns of prefrontal magnetic brain stimulation. Hum Brain Mapp. 2018;39(11):4580–4592.

27. Ahdab R, Ayache SS, Brugieres P, Goujon C, Lefaucheur JP. Comparison of “standard” and “navigated” procedures of TMS coil positioning over motor, premotor and prefrontal targets in patients with chronic pain and depression. Neurophysiol Clin. 2010;40(1):27–36.

28. Fitzgerald PB, Hoy KE, Herring SE, et al. A double blind randomized trial of unilateral left and bilateral prefrontal cortex transcranial magnetic stimulation in treatment resistant major depression. J Affect Disord. 2012;139(2):193–198.

29. Li CT, Cheng CM, Chen MH, et al. Antidepressant Efficacy of Prolonged Intermittent Theta Burst Stimulation Monotherapy for Recurrent Depression and Comparison of Methods for Coil Positioning: A Randomized, Double-Blind, Sham-Controlled Study. Biol Psychiatry. 2020;87(5):443–450.

30. Blumberger DM, Maller JJ, Thomson L, et al. Unilateral and bilateral MRI-targeted repetitive transcranial magnetic stimulation for treatment-resistant depression: a randomized controlled study. J Psychiatry Neurosci. 2016;41(4):E58–66.

31. Paillere Martinot ML, Galinowski A, Ringuenet D, et al. Influence of prefrontal target region on the efficacy of repetitive transcranial magnetic stimulation in patients with medication-resistant depression: a [(18)F]-fluorodeoxyglucose PET and MRI study. Int J Neuropsychopharmacol. 2010;13(1):45–59.

32. Moher D, Liberati A, Tetzlaff J, Altman DG. Preferred reporting items for systematic reviews and meta-analyses: the PRISMA statement. Int J Surg. 2010;8(5):336–341.

33. Moher D, Liberati A, Tetzlaff J, Altman DG, Group P. Preferred reporting items for systematic reviews and meta-analyses: the PRISMA statement. PLoS medicine. 2009;6(7):e1000097.

34. Li C-T, Cheng C-M, Chen M-H, et al. Antidepressant efficacy of prolonged intermittent theta burst stimulation monotherapy for recurrent depression and comparison of methods for coil positioning: a randomized, double-blind, sham-controlled study. Biological psychiatry. 2020;87(5):443–450.

35. Blumberger DM, Maller JJ, Thomson L, et al. Unilateral and bilateral MRI-targeted repetitive transcranial magnetic stimulation for treatment-resistant depression: a randomized controlled study. Journal of psychiatry & neuroscience: JPN. 2016;41(4):E58.

36. Miller IW, Bishop S, Norman WH, Maddever H. The modified Hamilton rating scale for depression: reliability and validity. Psychiatry research. 1985;14(2):131–142.

37. Zheng Y, Zhao J, Phillips M, et al. Validity and reliability of the Chinese Hamilton depression rating scale. The British Journal of Psychiatry. 1988;152(5):660–664.

38. Kühner C, Bürger C, Keller F, Hautzinger M. Reliability and validity of the revised Beck Depression Inventory (BDI-II). Results from German samples. Der Nervenarzt. 2007;78(6):651–656.

39. Zhang Y, Wang Y, Qian M. Reliability and validity of Beck Depression Inventory (BDI) examined in Chinese samples. Chinese Mental Health Journal. 1990;4(4):22–26.

40. Davidson J, Turnbull CD, Strickland R, Miller R, Graves K. The Montgomery - Absberg Depression Scale: reliability and validity. Acta psychiatrica scandinavica. 1986;73(5):544–548.

41. Drieling T, Schärer L, Langosch J. The Inventory of Depressive Symptomatology: German translation and psychometric validation. International journal of methods in psychiatric research. 2007;16(4):230–236.

42. Furukawa TA, Reijnders M, Kishimoto S, et al. Translating the BDI and BDI-II into the HAMD and vice versa with equipercentile linking. Epidemiology and psychiatric sciences. 2020;29.

43. Leucht S, Fennema H, Engel RR, Kaspers-Janssen M, Szegedi A. Translating the HAM-D into the MADRS and vice versa with equipercentile linking. Journal of affective disorders. 2018;226:326–331.

44. Sonmez AI, Camsari DD, Nandakumar AL, et al. Accelerated TMS for depression: a systematic review and meta-analysis. Psychiatry research. 2019;273:770–781.

45. Bais L, Vercammen A, Stewart R, et al. Short and long term effects of left and bilateral repetitive transcranial magnetic stimulation in schizophrenia patients with auditory verbal hallucinations: a randomized controlled trial. PloS one. 2014;9(10):e108828.

46. Loo CK, Sainsbury K, Mitchell P, Hadzi-Pavlovic D, Sachdev PS. A sham-controlled trial of left and right temporal rTMS for the treatment of auditory hallucinations. Psychological medicine. 2010;40(4):541–546.

47. Higgins JP, Thomas J, Chandler J, et al. Cochrane handbook for systematic reviews of interventions. John Wiley & Sons; 2019.

48. Hedges LV, Olkin I. Statistical methods for meta-analysis. Academic press; 2014.

49. Higgins JP, Thompson SG, Deeks JJ, Altman DG. Measuring inconsistency in meta-analyses. Bmj. 2003;327(7414):557–560.

50. Berlim M, Van den Eynde F, Daskalakis ZJ. A systematic review and meta-analysis on the efficacy and acceptability of bilateral repetitive transcranial magnetic stimulation (rTMS) for treating major depression. Psychological medicine. 2013;43(11):2245–2254.

51. Bowden J, Tierney JF, Copas AJ, Burdett S. Quantifying, displaying and accounting for heterogeneity in the meta-analysis of RCTs using standard and generalised Q statistics. BMC medical research methodology. 2011;11(1):1–12.

52. Che X, Cash R, Chung S, Fitzgerald PB, Fitzgibbon BM. Investigating the influence of social support on experimental pain and related physiological arousal: A systematic review and meta-analysis. Neuroscience & Biobehavioral Reviews. 2018;92:437–452.

53. Qin X-Y, Feng J-C, Cao C, Wu H-T, Loh YP, Cheng Y. Association of peripheral blood levels of brain-derived neurotrophic factor with autism spectrum disorder in children: a systematic review and meta-analysis. JAMA pediatrics. 2016;170(11):1079–1086.

54. Thompson SG, Higgins JP. How should meta - regression analyses be undertaken and interpreted? Statistics in medicine. 2002;21(11):1559–1573.

55. Van Houwelingen HC, Arends LR, Stijnen T. Advanced methods in meta - analysis: multivariate approach and meta-regression. Statistics in medicine. 2002;21(4):589–624.

56. Stanley TD, Jarrell SB. Meta - regression analysis: a quantitative method of literature surveys. Journal of economic surveys. 2005;19(3):299–308.

57. Light R, Pillemer D. Summing up: The science of research reviewing. In: Cambridge, MA: Harvard University Press; 1984.

58. Sterne JA, Sutton AJ, Ioannidis JP, et al. Recommendations for examining and interpreting funnel plot asymmetry in meta-analyses of randomised controlled trials. Bmj. 2011;343.

59. Egger M, Smith GD, Schneider M, Minder C. Bias in meta-analysis detected by a simple, graphical test. Bmj. 1997;315(7109):629–634.

60. Higgins JP, Savović J, Page MJ, Elbers RG, Sterne JA. Assessing risk of bias in a randomized trial. Cochrane handbook for systematic reviews of interventions. 2019:205–228.

61. Lin L, Chu H. Quantifying publication bias in meta-analysis. Biometrics. 2018;74(3):785–794.

62. Wood L, Williams C, Billings J, Johnson S. A systematic review and meta-analysis of cognitive behavioural informed psychological interventions for psychiatric inpatients with psychosis. Schizophrenia Research. 2020;222:133–144.

63. Kennedy NI, Lee WH, Frangou S. Efficacy of non-invasive brain stimulation on the symptom dimensions of schizophrenia: a meta-analysis of randomized controlled trials. Eur Psychiatry. 2018;49:69–77.

64. Shen X, Liu M, Cheng Y, et al. Repetitive transcranial magnetic stimulation for the treatment of post-stroke depression: a systematic review and meta-analysis of randomized controlled clinical trials. J Affect Disord. 2017;211:65–74.

65. Price GW, Lee JW, Garvey C-AL, Gibson N. The use of background EEG activity to determine stimulus timing as a means of improving rTMS efficacy in the treatment of depression: a controlled comparison with standard techniques. Brain stimulation. 2010;3(3):140–152.

66. Jha S, Chadda RK, Kumar N, Bal C. Brain SPECT guided repetitive transcranial magnetic stimulation (rTMS) in treatment resistant major depressive disorder. Asian journal of psychiatry. 2016;21:1–6.

67. Martinot M-LP, Galinowski A, Ringuenet D, et al. Influence of prefrontal target region on the efficacy of repetitive transcranial magnetic stimulation in patients with medication-resistant depression: a [18 F]-fluorodeoxyglucose PET and MRI study. The The International Journal of Neuropsychopharmacology. 2010;13(1):45–59.

68. Zhang Z, Zhang H, Xie C-M, et al. Task-related functional magnetic resonance imaging-based neuronavigation for the treatment of depression by individualized repetitive transcranial magnetic stimulation of the visual cortex. Science China Life Sciences. 2021;64(1):96–106.

69. Padberg F, Zwanzger P, Keck ME, et al. Repetitive transcranial magnetic stimulation (rTMS) in major depression: relation between efficacy and stimulation intensity. Neuropsychopharmacology. 2002;27(4):638–645.

70. Wang JX, Rogers LM, Gross EZ, et al. Targeted enhancement of cortical-hippocampal brain networks and associative memory. Science. 2014;345(6200):1054–1057.

71. Rigucci S, Serafini G, Pompili M, Kotzalidis GD, Tatarelli R. Anatomical and functional correlates in major depressive disorder: the contribution of neuroimaging studies. World J Biol Psychiatry. 2010;11(2 Pt 2):165–180.

72. Kimbrell TA, Little JT, Dunn RT, et al. Frequency dependence of antidepressant response to left prefrontal repetitive transcranial magnetic stimulation (rTMS) as a function of baseline cerebral glucose metabolism. Biol Psychiatry. 1999;46(12):1603–1613.

73. Loo CK, Sainsbury K, Mitchell P, Hadzi-Pavlovic D, Sachdev PS. A sham-controlled trial of left and right temporal rTMS for the treatment of auditory hallucinations. Psychol Med. 2010;40(4):541–546.

74. Blumberger DM, Mulsant BH, Fitzgerald PB, et al. A randomized double-blind sham-controlled comparison of unilateral and bilateral repetitive transcranial magnetic stimulation for treatment-resistant major depression. World J Biol Psychiatry. 2012;13(6):423–435.

75. Fitzgerald PB, Benitez J, de Castella A, Daskalakis ZJ, Brown TL, Kulkarni J. A randomized, controlled trial of sequential bilateral repetitive transcranial magnetic stimulation for treatment-resistant depression. Am J Psychiatry. 2006;163(1):88–94.

76. Medaglia JD, Lynall ME, Bassett DS. Cognitive network neuroscience. Journal of cognitive neuroscience. 2015;27(8):1471–1491.

77. Supekar K, Musen M, Menon V. Development of large-scale functional brain networks in children. PLoS Biol. 2009;7(7):e1000157.

78. Aguirre I, Carretero B, Ibarra O, et al. Age predicts low-frequency transcranial magnetic stimulation efficacy in major depression. J Affect Disord. 2011;130(3):466–469.

79. Begemann MJ, Brand BA, Curcic-Blake B, Aleman A, Sommer IE. Efficacy of non-invasive brain stimulation on cognitive functioning in brain disorders: a meta-analysis. Psychol Med. 2020;50(15):2465–2486.

